# Quantitative and qualitative effects of live music medicine on anxiety and depression in cancer inpatients

**DOI:** 10.1101/2022.10.19.22281272

**Authors:** Michael Rosenheck, Robert Hirsh

## Abstract

**Background:** A growing body of literature suggests that music medicine may decrease anxiety and depression in cancer patients, but the mechanism by which this happens remains speculative. This study analyzes the underlying thematic perspectives by utilizing both quantitative and qualitative assessments. Therefore, the goal of this study is to determine the quantitative effects and underlying thematic perspectives of music medicine on anxiety and depression in cancer inpatients.

**Methods and findings:** Twenty-four cancer inpatients participated in this prospective cohort study to measure the effects of a private, fifteen-minute, live guitar/singing performance on anxiety and depression. Questions modified from the Hospital Anxiety and Depression Scale (HADS) and a written free response section were used. Independent from age, gender, and relationship to music, music medicine significantly increased patients’ cheerfulness, ability to laugh, relaxation, and decreased worrying thoughts. There was also a trend towards decreased tension, restlessness, feeling slowed down, and increased excitement for the future that failed to reach statistical significance. The most frequently used words within the free response sections were made into a word cloud with the three most common words being music, listening, and made.

**Conclusion:** This article not only illustrates that music medicine decreases several elements of anxiety and depression in cancer inpatients, but also highlights music’s physiological effects, aesthetic and potentially transcendent properties, intrinsic value, and memorability, through use of a word cloud. Music medicine is a safe and inexpensive mood augmenter that could be more widely used.

## Introduction

Patients with cancer often experience emotional, physical, and social suffering. Between 30 and 50% of patients have moderate to high level of distress during the course of their illness[1, 2]. Studies have identified many negative consequences that are correlated with distress in cancer patients, including decreased quality of life[3], longer hospital stays[4], decreased treatment compliance[5], higher rates of suicide[6], and decreased survival[7]. Therefore, addressing cancer patient distress is of the upmost importance to achieve successful outcomes.

Music is one of the most used strategies to cope with illness[8], and music interventions have been used in several capacities throughout a cancer patient’s hospital course[9]. A recent systematic review showed that musical intervention is correlated with improved anxiety, pain, fatigue, and quality of life in people with cancer[10]. Many studies have shown the beneficial effects in reducing anxiety and depression, perception of pain, and physiologic markers of stress, including heart rate, respiratory rate, blood pressure[11], cortisol, and blood glucose[12].

The use of music in a clinical setting can be classified under two different umbrellas: music therapy and music medicine[13]. Music therapy involves a trained music therapist’s relationship with a patient. The music therapist provides an intervention based upon an assessment and evaluation of the needs of the patient. In contrast, music medicine is a more passive musical experience in which a patient listens to music. There is not a therapeutic relationship between a musician and a patient, and therefore no assessments nor treatments. Interestingly, systematic review found music therapy and music medicine equally effective in improving psychological outcomes in cancer patients[14].

Toccafondi et al. sought to evaluate the impact of a single-session musical medicine intervention on the wellbeing of cancer inpatients[9]. The intervention consisted of a classical music performance by a group of musicians called “Music Givers”, followed by a dinner buffet. They found that compared to patients who did not attend the concert, those who did attend demonstrated less distress at discharge and decreased symptoms of anxiety and depression. The authors of this study postulated two possible explanations for their positive findings. First, listening to music in a group setting may establish relationships between cancer inpatients. Second, listening to music may serve as a refuge from the worries related to illness. However, the dinner buffet may have confounded the positive findings.

The current study further explores the thematic perspectives by which music medicine impacts cancer inpatients and eliminates the dinner confounder found in the “Music Givers” format[9]. Additionally, this intervention is simpler to implement by not using licensed music therapists. During times of the Covid-19 pandemic and for other cancer patients, it is important that interactions between immunocompromised patients must be limited. Therefore, delivering music medicine in a group setting is undesirable. Consequently, the current study provides live music medicine in a private setting (within the patients’ rooms) – isolating the question, does music medicine improve the well-being of cancer inpatients, and if so, how?

This study utilizes both quantitative and qualitative data to improve understanding of how music medicine impacts patients. The hypothesis of this study is that a private music medicine format will decrease the anxiety and depression of cancer inpatients by virtue of thematic perspectives described by O’Callaghan et al. (2016) such as refuge from the stresses of their current illness.[15] Word clouds have been used recently for the visual representation of qualitative data[16-18] and this approach was chosen as a simple way of presenting the qualitative information. Word clouds are a visually exciting way to gain a basic understanding of qualitative data quickly and effectively[19, 20]. They are especially useful when analyzing preliminary data[21].

This study will help delineate whether isolated music medicine (delivered by a musician) in contrast to music therapy (delivered by a trained therapist) is beneficial to the patient, while using the patients’ own words to provide qualitative insight into their experiences.

In particular during this pandemic, the musical intervention proves to be a safe, inexpensive, and an effective stress-modifier that may lead to better health outcomes. Positive results argue for the wider use of music medicine in the inpatient setting.

## Materials and Methods

This preliminary study was designed to identify potentially positive outcomes from music in a way that is least obtrusive to the patient. If findings are positive, it would be anticipated that this study would be repeated with a control group and with the collections of comprehensive demographic data. The procedure used in this study did not require written informed consent.

### Ethics

The study was approved by the Institutional Review Board (IRB) and Human Research Protection Program (HRPP) of Cooper University Hospital and MD Anderson Cancer Center. This trial is registered as International Standard Randomized Controlled Trial Number 28383230. Each participant was given an information sheet which included the study title, names and phone numbers of the investigators, common questions and answers, research purpose, and risks and benefits of participation. Verbal consent was obtained from all participants. Additionally, word clouds are licensed under the Creative Commons License and can be used so long as it is not for profit and with proper citation.

### Study Sample

This quantitative and qualitative prospective cohort study included 24 participants and was carried out on the inpatient cancer floor of Cooper University Hospital and MD Anderson Cancer Center, Camden, New Jersey, USA from April to July of 2021. Cooper University Hospital serves an ethnically, racially, and socioeconomically diverse patient population.

Participation in the study was proposed to all cancer patients on the inpatient cancer floor, regardless of site or stage of disease. The exclusion criteria were: (1) those who are deaf (2) unable to read, write, or comprehend the survey (3) had previously participated in the study (4) under 18 years old (5) on end-of-life care or too critically ill to participate (6) those that have more than one patient per room (7) or if the clinical team did not authorize the encounter. Care was taken to avoid coercion to participate in the study, emphasizing that participation was voluntary, that their choice would not affect their medical care, and they would still be eligible to receive music medicine if they chose to abstain from the study.

### Study design

The study used one musician who was a male in his 20s. The musician had not previously met any participant. In the patient rooms, the musician used a standardized script for introduction, description of the study, and inquiry as to interest in study participation. After giving verbal consent, the participants received an information sheet about the study and were asked to complete the pre-intervention survey once the musician left the room. After one hour, the musician returned to the patient room to collect the pre-intervention survey and sing two standardized songs with acoustic guitar self-accompaniment. Then, the participants were asked to complete the post-intervention survey once the musician left the room. The post-intervention surveys were collected approximately fifteen minutes later.

### Data Collection

All data was collected via participant surveys: pre and post. The pre-intervention survey consisted of 13 multiple choice questions about age range, gender, anxiety, depression, and relationship to music. The questions about anxiety and depression were adapted and modified from The Hospital Anxiety and Depression Scale (HADS)[22]. These modifications include (1) decreasing number of items from 14 (7 on anxiety, 7 on depression) to 8 (4 on anxiety, 4 on depression), (2) shortening the length of some question stems, (3) transitioning from a 4-point to a 7-point Likert scale to quantify subtle changes more precisely, and (4) altering items to measure participants’ current feelings rather than their feelings over the past week. The survey was shortened for this preliminary study to limit burden and survey fatigue in an already fatigued patient population. The pre- and post-intervention surveys can be found in appendices 1 and 2 respectively.

### Statistical analysis

Differences in levels of anxiety, depression, and relationship to music from pre-intervention to post-intervention surveys were assessed using the Wilcoxon ranked sign test. SPSS was used. Effects of age, gender, and relationship to music were analyzed using ANOVA of normalized ranks. A word cloud was generated to analyze the free response section using Worditout.com. Only words used at least twice were included. Predetermined words commonly used in the English language, such as “a”, “it”, “the”, etc. were excluded.

## Results

### Description of the sample

A total of 180 inpatients were assessed for eligibility. 154 patients were excluded because they did not meet inclusion criteria (117), or they declined to participate (37). This left 26 participants. Out of this sample, 24 (92.3%) were able to fully complete the survey. Two participants did not complete the entirety of the surveys and where therefore excluded from analysis. A flow diagram depicting this can be found in Fig 1.

**Figure 1.**
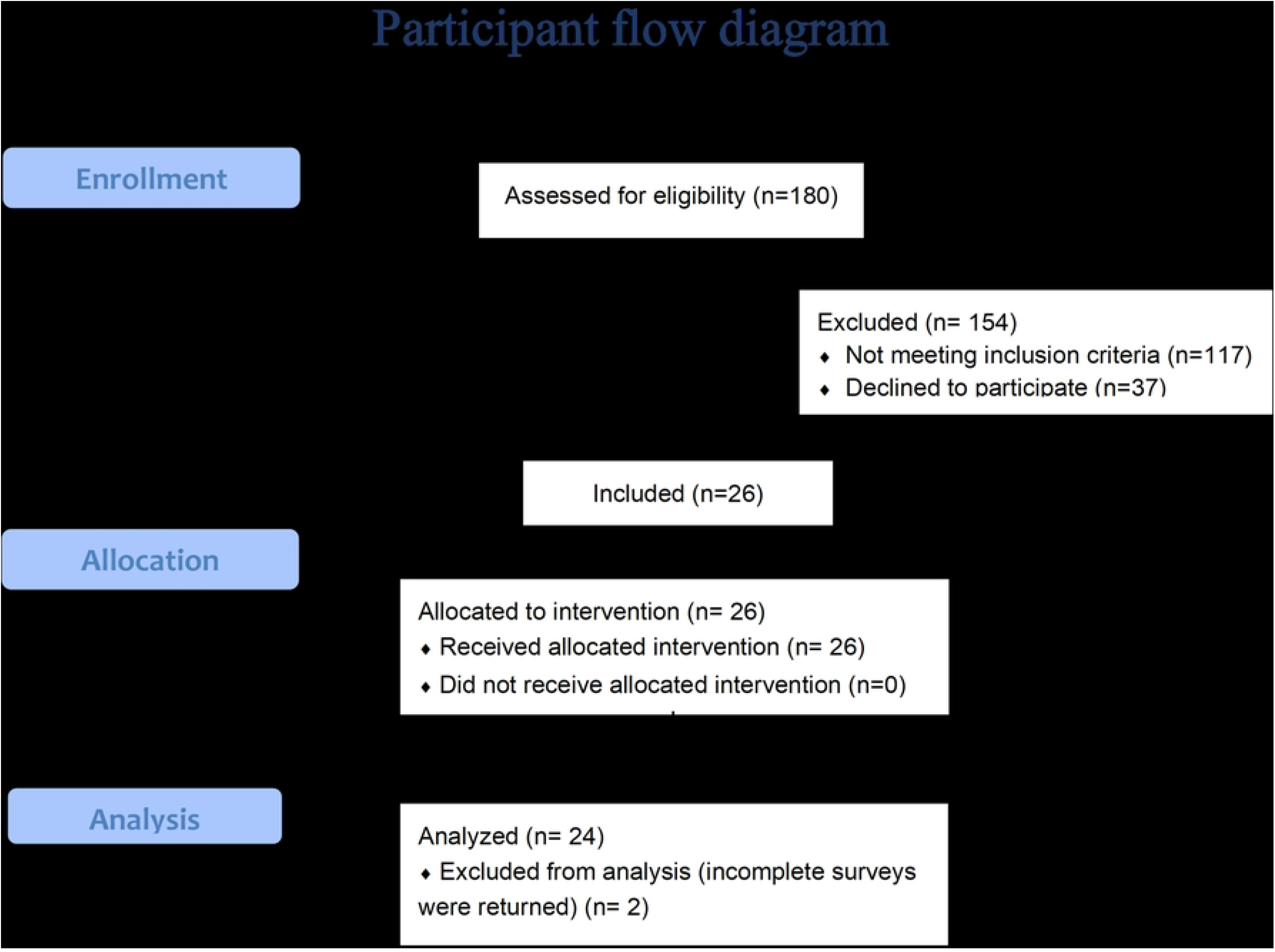
Flow diagram of number of participants from enrollment through analysis.

Patient demographics and music listening frequencies are summarized in Table 1.

**Table 1.** Description of basic participant demographics and frequency in which they listen to live music.

|  | Frequency<br>(N = 24) | Percent<br>% |
| --- | --- | --- |
| **Gender** |  |  |
| female | 17 | 70.83 |
| male | 7 | 29.17 |
| other | 0 | 0 |
| prefer not to say | 0 | 0 |
| **Age** |  |  |
| 18-30 | 0 | 0 |
| 31-40 | 4 | 16.67 |
| 41-50 | 4 | 16.67 |
| 51-60 | 6 | 25.00 |
| 61-70 | 7 | 29.17 |
| 71-80 | 3 | 12.50 |
| 81-90 | 0 | 0 |
| 91+ | 0 | 0 |
| **Frequency that patient listens to live music** |  |  |
| <1 time per 10 years | 4 | 16.67 |
| <1 time per year, >1 time per 10 years | 6 | 25.00 |
| 1-2 times per year | 4 | 16.67 |
| ≥3 times per year | 10 | 41.67 |

### Quantitative Effects of the Intervention on Anxiety and Depression

Analysis of pre- and post-intervention survey items relating to anxiety, depression, and relationship to music are summarized in Table 2.

**Table 2.** Analysis of survey items related to anxiety, depression, and relationship to music before verses after music intervention.

| Category | Question | pre-intervention<br>(n = 24) |  | post-intervention<br>(n = 24) |  | *p* – value* |
| --- | --- | --- | --- | --- | --- | --- |
|  |  | Mean | <u>SD</u> | <u>Mean</u> | <u>SD</u> |  |
| Anxiety | Tension | 3.71 | <u>1.90</u> | <u>3</u> | <u>2.06</u> | <u>0.177</u> |
| Anxiety | Worry thoughts | 4.46 | <u>2.04</u> | <u>2.88</u> | <u>1.73</u> | **<u>0.011</u>\*** |
| Anxiety | At ease/relaxed | 4.33 | <u>1.63</u> | <u>5.54</u> | <u>1.61</u> | **<u>0.008</u>\*** |
| Anxiety | Restlessness | 3.79 | <u>1.96</u> | <u>2.96</u> | <u>1.71</u> | <u>0.139</u> |
| Depression | Ability to laugh | 5 | <u>1.56</u> | <u>6</u> | <u>1.29</u> | **<u>0.013\*</u>** |
| Depression | Cheerfulness | 4.17 | <u>1.52</u> | <u>5.67</u> | <u>1.63</u> | **<u>0.002\*</u>** |
| Depression | Slowed down | 5.13 | <u>1.68</u> | <u>4.63</u> | <u>1.66</u> | <u>0.305</u> |
| Depression | Excitement for the future | 5.29 | <u>1.60</u> | <u>5.67</u> | <u>1.24</u> | <u>0.436</u> |
| Music | Like music | 6.46 | <u>1.06</u> | <u>6.54</u> | <u>0.98</u> | <u>0.602</u> |
| Music | Music large part of life | 5.67 | <u>1.58</u> | <u>6.08</u> | <u>1.32</u> | <u>0.277</u> |
| Music | Thought about other things | NA | <u>NA</u> | <u>6.58</u> | <u>0.72</u> | <u>NA</u> |

\**p* values were calculated using the Wilcoxon ranked sign test.

Through ANOVA of normalized ranks, there were no significant effects of sex, age range, or relationship to music on tension, ability to laugh, worry, cheerfulness, relaxation, feeling slowed down, restlessness, or excitement for the future (*p* > 0.05).

### Qualitative effects of Intervention

In response to the free response question on the post-intervention survey “please write in your own words how listening to music made you feel”, a word cloud from www.worditout.com was generated containing only words used at least twice, excluding predetermined words commonly used, such as “a”, “it”, “the”, etc. The three most often used words were “music” (eleven times), “listening” (seven times), and “made” (five times). Words used four times include “relaxed” and “relaxing”. Words used three times include “feel”, “back”, “like” and “life”. Words used two times include “thank”, “took”, “voice”, “dark”, “happy”, “soothing”, “brought”, “nice”, “take”, “just”, “felt”, “old”, “play”, “away”, “more”, “good”, “enjoyed”, “songs”, and “see”. For the word cloud representation, please see Fig 2.

**Figure 2.**
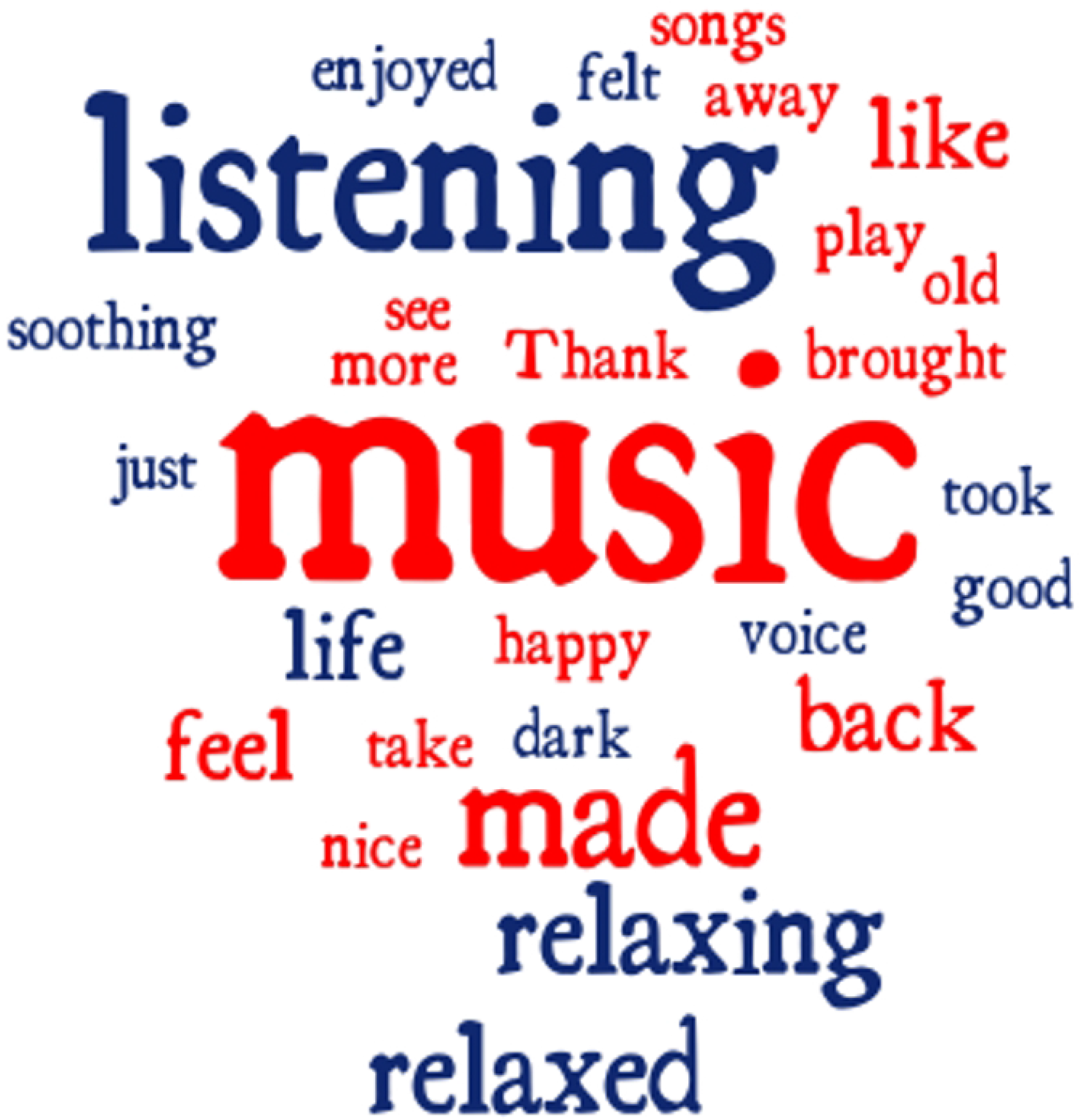
Word cloud of most frequently used words by patients when describing their music medicine experience[23].

There were no adverse effects of this study.

## Discussion

Music medicine decreased several aspects of anxiety and depression in cancer inpatients. Specifically, it decreased worrying thoughts and increased ease and relaxation, cheerfulness, and the ability to laugh. For the items that did not reach statistical significance, including “tension,” “restlessness,” “excitement for the future,” and “feeling slowed down,” there were clear trends towards decreased anxiety and depression. Failure to reach significance may have been due to small sample size.

Regarding anxiety, our findings were consistent with a 2017 metanalysis that used 9 studies[24]. They found that music significantly reduced anxiety in cancer patients. However, there was much heterogeneity in methodologies between selected studies, including active and passive listening, number of sessions, live and recorded music, and therapeutic and non-therapeutic relationships. The “Music Givers” format employed in Toccafondi et al 2017[9], is most like our study. Both used passive, one-time, live music medicine sessions performed by trained musicians rather than music therapists. Our studies differ in that the Music Givers format was performed in front of a crowd of cancer patients and their family members, whereas ours took place inside individualized patient rooms. Additionally, the Music Givers format included a free buffet following the performance, whereas we did not provide food. Using the HADS, they found that patients that attended the Music Givers performance experienced less anxiety upon hospital discharge compared to those who did not attend the performance. Our study was designed to avoid the possible confounder of food consumption.

There remains no consensus about music’s effect on depression in cancer patients in the literature. Two metanalyses revealed that music interventions significantly reduced depression in patients with cancer[25, 26]. While two separate meta-analyses did not find effects of music therapy or music medicine on depression in patients with cancer[11, 24]. These inconsistencies may be due to limited sample sizes and studies with less rigorous design, highlighting the need for further research. This study offers support that music medicine reduces report of symptoms associated with depression and anxiety in cancer inpatients.

Interestingly, the effects of music medicine do not differ based on gender, age, or music background. This speaks to music being a universal language that everyone can appreciate regardless of background.

By using the participants’ own words, the word cloud (Fig. 2) was generated for qualitative analysis. One approach to interpret the word cloud is to sort each word into categories based on their themes to gain better understanding of mechanisms by which music alters emotion in cancer patients. Through a meta-ethnography of 138 participants, O’Callaghan et al. described nine thematic perspectives by which cancer patients experienced music[15]. Five of these themes were reflected in our word cloud: the physical effects of music, the aesthetic and potentially transcendent effects of music, the intrinsic value of music, the common ontological qualities between music and cancer, and the reminiscent effects of music. The four themes not reflected in our word cloud include music as a form of communication, music as a means of creativity, music as symbolism, and the effects of music on accommodating projections and self-identifications[15]. These last four themes were not seen due to the passive nature of music medicine in comparison to music therapy where the patient often takes an active role in the musical process. These results are summarized in Table 3.

**Table 3.** Most used words sorted into thematic perspectives as characterized in O’Callaghan et al. 2016[15].

| Thematic Perspectives (O’Callaghan) | Words |
| --- | --- |
| Music Has Physiological Effects. | enjoyed, like, good, happy, nice, made, felt, feel |
| Music Is Aesthetic and a Potentially Transcendent Experience. | brought, took, take, away, back, old |
| Music Can Be a Transitional Phenomenon and Means to Meaningfully Communicate. | na |
| Music Is Intrinsically Valuable. | enjoyed, like, good, happy, nice, made, felt, feel |
| Music Can Be Symbolic. | na |
| Musical Experience Is Creative. | na |
| Music Can Accommodate Projections and Identifications. | na |
| Music Shares Ontological Properties with Cancer. | dark |
| Music Is Remembered. | brought, took, take, away, back, old |

Words such as *enjoyed, like, good, happy, nice, made, felt*, and *feel* speak to music’s intrinsic value as well as its physiological effects. These words are involved in the neural activation of arousal and pleasure [27] and therefore cause physiological change. Words like *brought, took, take, away, back*, and *old* are consistent with the transcendent and reminiscent effects of music. When listening to music, some cancer patients are momentarily transported into a different headspace, often involving a comforting memory. This can be relieving for patients whose lives have been overrun by cancer. The word *dark* may reflect the shared ontological qualities between music and cancer. Cancer can symbolically be represented by music because they both may be elusive, unpredictable, and mysterious. Additionally, cancer is life threatening and music is transient in nature. Therefore, music can touch, understand, and alter cancer’s effects, offering hope to patients[28].

The remaining words on the word map are more related to the performance than the patient’s emotional reactions. These words include *songs, listening, play, music, voice, see, more*, and *just*.

This study has limitations. The sample size was small in this preliminary study. Further, the surveys were modified from the HADS, and the instrument used cannot be considered as previously validated. This study only analyzed patients who wanted music medicine and excluded those who declined it. Therefore, the study did not have a control group. Since this study was designed to be introductory, its purpose was to identify potential positive findings. In summary, a future study would include written informed consent, collection of more robust demographic data, data collection including a control group, full use of a validated survey, and a larger sample size. Additionally, since this study excluded those who declined music medicine, the findings are not generalizable to the public, but rather to those who are open to receiving music medicine.

## Conclusion

Music medicine proved to be an effective way of decreasing reported symptoms of anxiety and depression in cancer inpatients during the COVID-19 pandemic when family and friend visitation was limited. Specifically, live music medicine significantly increased relaxation, cheerfulness, and ability to laugh, and significantly decreased worrying thoughts. This warrants more robust research to determine whether the other symptoms that trended towards improvement will significantly improve with a larger sample size. Additionally, through use of the word cloud, this study further uncovered the thematic perspectives by which music effects cancer inpatients, warranting replication studies.

## Data Availability

All data may be found in the attached spreadsheet labeled "S1 supporting information".

## Acknowledgements

The study team would like to acknowledge Susan Cavanaugh, Dr. Faith Young, Dr. Danielle Behrens, and Dr. Lori Winter-Feldman for their collaboration, insight, and support, Dr. John Gaughan for his help with statistical analysis, Dr. David and Rebecca Rosenheck, Dr. Eve Rosenheck, Dr. Paul Wallach, and Miriam Wallach for their help editing the manuscript, and Heather Brown and the rest of the nursing staff on 5^th^ floor Pavilion for their support, positivity, and excellent care.

## Supporting information

**S1 Appendix. Sample pre-intervention survey given to all participants one hour before music intervention**.

**S2 Appendix. Sample post-intervention survey given to all participants immediately after music intervention**.

**S1 Supporting Information. Raw data set of survey responses**.

**S2 Supporting Information. IRB protocol**.

**S3 Supporting Information. TREND Checklist**.

## References

1. Carlson L, Angen M, Cullum J, Goodey E, Koopmans J, Lamont L, et al. High levels of untreated distress and fatigue in cancer patients. British journal of cancer. 2004;90(12):2297–304.

2. Zabora J, BrintzenhofeSzoc K, Curbow B, Hooker C, Piantadosi S. The prevalence of psychological distress by cancer site. Psycho-Oncology: Journal of the Psychological, Social and Behavioral Dimensions of Cancer. 2001;10(1):19–28.

3. Brown LF, Kroenke, K., Theobald, D.E., et al. The assocaition of depression and anxiety with health-related quality of life in cancer patients with depression and/or pain. Psycho-Oncology. 2010:734–41.

4. Prieto JM, Blanch, J., et al. Psychiatric morbidity and impact on hospital length of stay among hematologic cancer patients recieving stem-cell transplantation. Journal of Clinical Oncology. 2002:1907–17.

5. Berry DL, Blonquist, T.M., Hong, F., et al. Self-reported adherence to oral cancer therapy: Relationaships with symptom distress, depression, and personal characteristics. Patient Preference and Adherence. 2015:1587–92.

6. Breitbart W, Rosenfeld, B., Pessin, H., et al. Depression, hopelessness, and desire for hastened death in terminally ill patients with cancer. The Journal of the American Medical Association. 2000:2907–11.

7. Kissane D. Beyond the psychotherapy and survival debate: The challenge of social disparity, depression and treatment adherence in psychosocial cancer care. Psycho-Oncology. 2009:1–5.

8. Zaza C, Sellick, S.M. & Hillier, L.M. Coping with cancer: What do patients do?. Journal of Psychosocial Oncology. 2005:55–73.

9. Toccafondi A, Bonacchi, A., et al.,. Live music intervention for cancer inpatients: The Music Givers format Palliative and Supportive Care 2017:777–84.

10. Bradt J DC, Magil L, Teague A. Music interventions for improving psychological and physical outcomes in cancer patients Cochrane database of systematic reviews. 2016.

11. Bradt J, Dileo, C., Grocke, D., et al. Music interventions for improving psychological and physical outcomes in cancer patients.. The Cochrane Database for Systematic Reviews. 2011.

12. Finn S FD. The biological impact of listening to music in clinical and nonclinical settings: a systematic review Progress in brain research,. 2018.

13. Dileo C, editor Music Therapy and Medicine: Theoretical and Clinical Applications. 1999; Silver Spring, MD: American Music Therapy Association.

14. Bradt J, Patvin, N., Kesslick, A., Shim, M., Radl, D., Schriver, E., Gracely, E., Komarnicky-Kocher, L.The impact of music therapy versus music medicine on psychological outcomes and pain in cancer patients: a mixed methods study. Support Care Cancer 2015:1261–71.

15. O’Callaghan CC MF, Reid P, Michael N, Hudson P, Zalcberg JR, Edwards J. Music’s Relevance for People Affected by Cancer: A Meta-Ethnography and Implications for Music Therapists. Journal of Music Therapy. 2016:398–429.

16. Vrain E, Lovett A. Using word clouds to present farmers’ perceptions of advisory services on pollution mitigation measures. Journal of Environmental Planning and Management. 2020;63(6):1132–49.

17. DeNoyelles A, Reyes-Foster B. Using word clouds in online discussions to support critical thinking and engagement. Online Learning. 2015;19(4):4.

18. Brooks B, Gilbuena D, Krause S, Koretsky M. Using word clouds for fast, formative assessment of students’ short written responses. Chemical Engineering Education. 2014;48(4):190–8.

19. Mathews D, Franzen-Castle L, Colby S, Kattelmann K, Olfert M, White A. Use of word clouds as a novel approach for analysis and presentation of qualitative data for program evaluation. Journal of Nutrition Education and Behavior. 2015;47(4):S26.

20. Edyburn DL. Word Clouds: Valuable Tools When You Can’t See the Ideas Through the Words. Journal of Special Education Technology. 2010;25(2):68.

21. McNaught C, Lam P. Using Wordle as a supplementary research tool. Qualitative Report. 2010;15(3):630–43.

22. Zigmond ASS, R.P. The Hospital Anxiety and Depression Scale.. Acta Psychiatrica Scandinavica,. 1983:361–70.

23. WordItOut. [cited 2022. Available from: https://worditout.com.

24. Bro ML JK, Hansen JB, Vuust P, Abildgaard N, Gram J, Johansen C. Kind of blue: A systematic review and meta-analysis of music interventions in cancer treatment.. Psychooncology 2018:386–400.

25. Tsai HF, Chen YR, Chung MH, Liao YM, Chi MJ, Chang CC, et al. Effectiveness of music intervention in ameliorating cancer patients’ anxiety, depression, pain, and fatigue: A meta-analysis. Cancer nursing. 2014;37(6):E35–E50.

26. Zhang J-M, Wang P, Yao J-x, Zhao L, Davis MP, Walsh D, et al. Music interventions for psychological and physical outcomes in cancer: a systematic review and meta-analysis. Supportive Care in Cancer. 2012;20(12):3043–53.

27. Levitin DJ. This is your brain on music. London: Atlantic; 2006.

28. Stein A. Music, mourning, and consolation.. Journal of the American Psychoanalytic Association. 783–811:52.

